# Is amnestic mild cognitive impairment a neuro-immune condition?

**DOI:** 10.1101/2024.05.05.24306882

**Authors:** Vinh-Long Tran-Chi, Michael Maes, Gallayaporn Nantachai, Solaphat Hemrungrojn, Marco Solmi, Drozdstoy Stoyanov, Kristina Stoyanova, Chavit Tunvirachaisakul

**Affiliations:** Ph.D. Program in Clinical Sciences, School of Global Health, Faculty of Medicine, Chulalongkorn University, Bangkok, Thailand; Department of Psychiatry, Faculty of Medicine, Chulalongkorn University, Bangkok, Thailand; Sichuan Provincial Center for Mental Health, Sichuan Provincial People’s Hospital, School of Medicine, University of Electronic Science and Technology of China, Chengdu, China; Key Laboratory of Psychosomatic Medicine, Chinese Academy of Medical Sciences, Chengdu, China; Research Institute, Medical University of Plovdiv, Plovdiv, Bulgaria; Department of Psychiatry, Medical University of Plovdiv, Plovdiv, Bulgaria; Kyung Hee University, Dongdaemun-gu, Seoul, South Korea; Cognitive Impairment and Dementia Research Unit, Department of Psychiatry, Faculty of Medicine, Chulalongkorn University, Bangkok, Thailand; Cognitive Fitness and Biopsychiatry Technology Research Unit, Faculty of Medicine, Chulalongkorn University, Bangkok, Thailand; Strategic Research and Innovation Program for the Development of MU - PLOVDIV–(SRIPD-MUP), European Union – NextGenerationEU; Somdet Phra Sungharaj Nyanasumvara Geriatric Hospital, Department of Medical Services, Ministry of Public Health, Chon Buri Province, Thailand; Department of Psychiatry, University of Ottawa, Ontario, Canada; Regional Centre for the Treatment of Eating Disorders and On Track, The Champlain First Episode Psychosis Program, Department of Mental Health, The Ottawa Hospital, Ontario, Canada; Ottawa Hospital Research Institute (OHRI), Clinical Epidemiology Program, University of Ottawa, Ottawa, Ontario, Canada; Department of Child and Adolescent Psychiatry, Charité Universitätsmedizin, Berlin, Germany

**Keywords:** Neuroimmune, inflammation, depression, neurocognition, immune biomarkers, chemokines

## Abstract

**Background:** The pathophysiology of amnestic Mild Cognitive Impairment (aMCI) is largely unknown, although some papers found signs of immune activation.

**Aims:** To assess the cytokine network in aMCI after excluding patients with major depression (MDD) and to examine the immune profiles of quantitative aMCI (qMCI) and distress symptoms of old age (DSOA) scores.

**Methods:** A cross-sectional study was conducted on 61 Thai aMCI participants and 60 healthy old adults (both without MDD). The Bio-Plex Pro human cytokine 27-plex test kit and LUMINEX 200 were used to assay cytokines/chemokines/growth factors in fasting plasma samples.

**Results:** aMCI is characterized by significant general immnosuppression, and reductions in T helper 1 (Th)1 and T cell growth profiles, the immune-inflammatory responses system, interleukin (IL)1β, IL6, IL7, IL12p70, IL13, GM-CSF, and MCP-1 as compared with controls. These 7 cytokines/chemokines exhibit neuroprotective effects at physiologic concentrations. In multivariate analyses, three neurotoxic chemokines, CCL11, CCL5, and CXCL8, emerged as significant predictors of aMCI. Logistic regression showed that aMCI was best predicted by combining IL7, IL1β, MCP-1, years of education (all inversely associated) and CCL5 (positively associated). We found that 38.2% of the variance in the qMCI score was explained by IL7, IL1β, MCP-1, IL13, years of education (inversely associated) and CCL5 (positively associated). The DSOA were not associated with any immune data.

**Discussion:** A dysbalance between lowered levels of neuroprotective cytokines and chemokines, and relative increases in neurotoxic chemokines are key factors in aMCI. Future MCI research should always control for the confounding effects of affective symptoms.

## Introduction

Among older adults, the occurrence of mild cognitive impairment (MCI) is quite significant, affecting approximately 10-15% of individuals aged 65 and above (Anderson, 2019). Amnestic MCI (aMCI) is characterized by mild deficits in various cognitive domains, such as episodic memory, executive functions, visuospatial skills, processing speed, cognitive flexibility, and problem-solving ability (Nantachai et al., 2023; Tran-Chi et al., 2024; Tunvirachaisakul et al., 2018). Nevertheless, individuals with aMCI generally demonstrate intact abilities related to everyday tasks, as observed in various studies (Dwolatzky et al., 2004; Gualtieri & Johnson, 2005; Hemrungrojn et al., 2021). aMCI is a cognitive stage that falls between the normal aging process and the onset of dementia (Anderson, 2019). It is worth noting that the annual conversion rate from aMCI to Alzheimer’s disease (AD) is approximately 16.5%. Nevertheless, a small percentage of patients with aMCI (8%) exhibit a remission from this condition, as noted by Petersen et al. (2010).

The evaluation of aMCI poses two significant challenges. At first, it is uncertain if the findings regarding the neurocognitive aspects of MCI in older individuals are still applicable when patients with major depression (MDD) are excluded. Depressive symptoms can significantly affect the neurocognitive aspects of MCI, which poses challenges in interpreting assessments conducted in MCI studies that did not exclude patients with MDD (Tran-Chi et al., 2024). In addition, it has been argued that the existing diagnostic criteria for aMCI are excessively permissive, as suggested by Maes and Tangwongchai (2021). The ongoing debate revolves around the distinctiveness of aMCI as a phenotype and the possibility that some individuals classified as aMCI could actually be part of the normal control sample (Maes & Tangwongchai, 2021; Tunvirachaisakul et al., 2018). In a study conducted by Tran-Chi et al. (2024), it was discovered that older individuals without MDD exhibited two distinct symptom dimensions. The first dimension, distress symptoms of old age (DSOA), encompasses emotions such as depression, anxiety, tension, and neuroticism. This aspect demonstrates a significant correlation with negative life events and adverse childhood experiences (Tran-Chi et al., 2024). The second dimension involves a quantitative score of aMCI severity (labeled as qMCI), which provides an indication of the extent of objective cognitive decline. The score is calculated by taking into account the first principal component of the Montreal Cognitive Assessment (MoCA) and the Mini Mental State Examination (MMSE) scores, as well as the modified Clinical Dementia Rating (CDR) (Tran-Chi et al., 2024). In addition, through cluster analysis, it has been determined that the diagnostic criteria for aMCI proposed by Petersen (2004) are excessively broad. This is due to the fact that the aMCI group consists of individuals with subjective indicators of cognitive impairment, specifically subjects with DSOA after removing those with MDD (Tran-Chi et al., 2024).

Research suggests that the activation of the immune system in the peripheral blood may contribute to neuroinflammation, thereby contributing to neurocognitive deficits and Alzheimer’s disease (Maes et al., 1999; Xie et al., 2022). Peripheral inflammatory responses and heightened microglia-associated signaling are now considered crucial phenomena in Alzheimer’s disease, as highlighted by Kinney et al. (2018). Additionally, some studies have indicated that MCI is linked to elevated levels of immune compounds such as interleukin-6 (IL-6) and C-reactive protein (CRP) in serum or cerebro-spinal fluid (CSF) (Schmidt-Morgenroth et al., 2023; Miaoyan Zheng et al., 2019). Interestingly, increased IL-6 and tumor necrosis factor (TNF)-α, have been linked to a higher likelihood of MCI progressing to Alzheimer’s disease (Contreras et al., 2022). A meta-analysis conducted by Shen et al. (2018) found that individuals with MCI have elevated levels of IL-6, MCP-1, and soluble TNF receptor 2 (sTNFR2) in their serum, in comparison to the control group. Nevertheless, another meta-analysis, conducted by Saleem et al. (2015), found no significant alterations in immune variables in individuals with MCI. This includes acute phase reactants, immunoglobulins, cytokines, chemokines, and adhesion molecules.

Nevertheless, there has been a lack of research investigating comprehensive cytokine/chemokine profiles in individuals with aMCI after excluding those with MDD. Additionally, there is a dearth of research exploring the connections between the quantitative qMCI and DSOA scores and the comprehensive immune profiles in those MCI subjects without MDD. It should be stressed that MDD is currently recognized as a neuro-immune disorder, marked by elevated levels of pro-inflammatory cytokines, acute phase proteins, and complement factors (Maes et al., 1990; Maes & Carvalho, 2018; Maes, 1993). Moreover, the heightened neurotoxicity resulting from the impact of pro-inflammatory cytokines/chemokines on neurons is linked to MDD as well as the intensity of depression and anxiety (Maes & Carvalho, 2018; Maes et al., 2023).

Thus, the aim of this study is to assess the cytokine/chemokine network in individuals who have aMCI but do not have MDD, as well as the immune profiles of the qMCI and DSOA scores. The specific hypotheses aim to uncover the presence of immune activation in individuals with aMCI, as indicated by their immune profiles. Additionally, activated immune profiles, including the macrophage M1 profile, are expected to predict the qMCI and DSOA scores.

## Methods

### Participants

A cross-sectional study was conducted to compare individuals with aMCI to a group of healthy control subjects. The study sample comprised individuals of both genders, with an age range spanning from 60 to 75 years. The study recruited healthy participants from Bangkok, Thailand, whereas individuals with aMCI were recruited from the Outpatient Department of the Dementia Clinic at King Chulalongkorn Memorial Hospital in Bangkok, Thailand from May 2022 to March 2023. The clinical Petersen’s criteria were utilized to diagnose aMCI in the older adult population. These criteria involve the identification of subjective and objective memory impairments, together with the lack of dementia and alterations in activities of daily living (ADL). Furthermore, people with amnestic mild cognitive impairment complied with the Petersen criteria and exhibited a modified Clinical Dementia Rating score of 0.5. The control group had a CDR score of 0 and did not meet Petersen’s criteria (Petersen, 2004). The healthy older adults were recruited from the Health Check-up Clinic, members of neighbourhoods’ senior clubs, and the healthy, elderly carers of individuals with aMCI) who were patients at the Dementia Clinic, and senior volunteers affiliated with the Red Cross.

Participants with stroke, Parkinson’s disease, any dementia subtype, multiple sclerosis, schizophrenia, bipolar disorder, autism, delirium, metabolic disorder, malaria, HIV, chronic obstructive pulmonary disease (COPD), chronic kidney disease, cancer, substance abuse, alcoholism, inability to speak or communicate, blindness or impaired vision even with corrective lenses, hearing loss, inability to sit stably due to physical conditions such as chronic pain or low back pain, and those who had undergone cognitive training within three months prior to the study were excluded from the study. Ultimately, the subjects were allocated to either of the two study groups, consisting of 60 individuals classified as healthy controls and 61 individuals diagnosed with aMCI.

Before taking part in the study, all controls and patients were required to submit written informed consent. The study conducted in this research adhered to ethical and privacy standards that are recognized both in Thailand and internationally. These standards are in accordance with the International Guideline for the Protection of Human Subjects, as mandated by influential documents such as the Declaration of Helsinki, the Belmont Report, the International Conference of Harmonization in Good Clinical Practice, and the CIOMS Guidelines. The present study received approval from the Institutional Review Board (IRB) of the Faculty of Medicine at Chulalongkorn University in Bangkok, Thailand (No. 0372/65).

### Clinical assessments

We used a semi-structured interview to collect socio-demographic data comprising age, sex, relationship status, year of education, and marital status. The scales used to assess cognition were the Thai Mini-Mental State Examination (MMSE) (Committee, 1993), the Thai Montreal Cognitive Assessment (MoCA) (Tangwongchai et al., 2009), and three rating scales of the Thai Consortium to Establish a Registry for Alzheimer’s Disease (CERAD) Neuropsychological Assessment Battery (Tunvirachaisakul et al., 2018). The MMSE is a 30[question assessment of cognitive function (Folstein et al., 1975). The Thai version of the Mini-Mental State Examination was developed in 1993 and has been extensively used in Thailand to screen cognitive impairment and dementia (Committee, 1993). The scale consists of six subtests measuring orientation, registration, attention, word recall, language, and computation. The total score ranges from 0 to 30. The MoCA was developed by Nasreddine et al. (2005) as an effective and applicable screening tool for cognitive disorders. The Thai version of MoCA was used to screen and monitor cognitive impairments in the clinical practice of neurocognitive disorders validated in the Thai setting by Tangwongchai et al. (2009). This test measures various cognitive domains, namely: visuospatial/executive, naming, attention, language, abstraction, delayed recall, and orientation. The total sum of all individual scores (out of 30 maximum possible points) represents the severity of cognitive impairment. The qMCI score was derived as the first principal component derived from the MoCA, MMSE, and the CDR score (Tran-Chi et al., 2024).

To evaluate distress, depression, and anxiety symptoms, we utilized various assessment tools. These included the State-Trait Anxiety Inventory (STAI) developed by Spielberger et al. (1971), the depression (HADS-D) and anxiety (HADS-A) subscales of the Thai version of Hospital Anxiety and Depression Scale developed by Nilchaikovit (1996), the Perceived Stress Scale (PSS) by Wongpakaran and Wongpakaran (2010), the neuroticism trait score from the Five Factor Model standardized psychometric pool of items (IPIP-NEO) by Yomaboot and Cooper (2016), and the Thai Geriatric Depression Scale (TGDS) by Yesavage (1983). In a study conducted by (Tran-Chi et al., 2024), the DSOA dimension was constructed through PCA using the first PC extracted from STAI, HADS-D, HADS-A, PSS, STAI, TGDS, and the neuroticism score. To obtain a more homogeneous cohort of patients with aMCI, we have excluded subjects with DSOA symptoms (n=9) as explained by (Tran-Chi et al., 2024). This selected and homogeneous aMCI subset (n=52) is known as mild cognitive dysfunction (mCoDy) to differentiate it from the broader MCI category (Tran-Chi et al., 2024)..

### Assays

To assay cytokines/chemokines we used fasting venous blood sampled between 7.00 a.m. and 9.00 a.m. We determined the concentrations of cytokines, chemokines, and growth factors in plasma samples using the 27-plex Bio-Plex Pro™ Human Chemokine Assays (Bio-Rad Laboratories, Inc. USA) as explained previously (Maes et al., 2023). The LUMINEX 200 apparatus was used to measure the fluorescence intensities (FI), which are more suitable than absolute concentrations (particularly when multiple plates are utilized). Therefore, we used the blank subtracted FI values in our statistical analysis. It was determined that the intra-assay coefficient of variation values for each analyte were all below 11.0%. The concentrations were ascertained utilizing the standard concentrations supplied by the manufacturer. Following this, the proportion of concentrations surpassing the minimum measurable concentration (OOR) was computed. The analytes are detailed in Electronic Supplementary File (ESF), Table 1, along with the percentage of values that surpassed the OORs. Approximations of values below OOR values were achieved by utilizing the sensitivity of the assays. In the statistical analysis conducted on individual cytokines, chemokines, and growth factors, analytes whose concentrations were not detectable (< lower than the OOR) in over 50% of all assays were not entered as IF values but as prevalences (dummy variables: measurable versus non-measurable). However, in case less than 20% of the analytes were measurable, the variables were excluded from the analyses. Following this, IL-2, IL-5, IL-15, IL-17, and VEGF were excluded from the analysis. ESF, Table 2 lists the variables utilized in the construction of the subsequent immune profiles. Where needed, cytokines were used after logarithmic (log10), square root, fractional rank-based normal transformations, or Winsorization.

### Data analysis

Differences in continuous variables between groups were checked using analysis of variance (ANOVA). Analysis of contingency tables (the χ2-test) was used to determine the association between nominal variables. Correlations between two variables were assessed using Pearson’s product-moment correlation coefficients. Multivariate and univariate general linear model (GLM) analysis was used to examine the relationships between diagnostic classifications and clinical and cognitive data after covarying for gender, age, and education. Subsequently, the estimated marginal means (SE) were computed from the GLM model after adjusting for the gender, age, and education variables. We performed multiple regression analysis (manual and automatic stepwise) to determine which test scores best predicted the clinical scores and to compute and display partial regression analysis. For this analysis, we always confirmed multivariate normality, homoscedasticity, and the absence of collinearity and multicollinearity. The study employed manual and automatic stepwise binary logistic regression analysis to evaluate the explanatory variables that significantly predicted the diagnosis of aMCI or mCoDy as the dependent variable. The control group served as the reference group. Odds ratios were calculated with 95% confidence intervals (CI), and Nagelkerke values were utilized to estimate the effect size. The regression analyses’ results were always bootstrapped using 1,000 bootstrap samples, and the latter were reported if the findings were not concordant. Statistical tests were two-tailed and a p-value of 0.05 was used for statistical significance. IBM SPSS Windows version 29 was used for all statistical analyses.

## Results

### Socio-demographic and clinical Data

**Table 1** displays the demographics and clinical features of the two research groups. The cognitive assessments demonstrated significant differences among the groups with lower MMSE, MoCA, and qMCI scores in aMCI than in controls. There was a trend towards increased DSOA scores in aMCI. Certain participants received treatment consisting of oral antidiabetica (6 controls and 10 aMCI, X^2^=1.08, df=1, p=0.299), antihypertensive drugs (17 controls and 26 aMCI, X^2^=2.70, df=1, p=0.101), and lipid-lowering drugs (36 controls and 36 aMCI, X^2^=0.12, df=1, p=0.912). Several of the participants consumed vitamin (A, B, C, D, E) preparations (34 controls and 30 aMCI, X^2^=0.68, df=1, p=0.409), fish oil (5 controls and 10 aMCI, X^2^=1.81, df=1, p=0.179), or folic acid (3 controls and 2 aMCI, Fisher’s exact probability test: p=0.680). However, no statistically significant correlations were observed between any of these medications and the immune profiles. Furthermore, the inclusion of these drug state variables as covariates in the GLM analyses had no impact on the outcomes detailed below. Consequently, the patients’ medication status had no bearing on the findings of this research.

### Differences in immune-inflammatory biomarkers scores between the diagnostic groups

**Table 2** indicates that there are significant associations between the diagnosis (aMCI/HC) and immune-inflammatory biomarkers after controlling for age, sex, and BMI. Sex (p=0.714), age (p=0.963), and BMI (p=0.0103) did not have any significant effect in the multivariate GLM analysis, whereas diagnosis yielded a significant effect (F=2.00, df=9/108, p=0.046, partial eta squared=0.143). Univariate GLM showed significant differences in Th1, IRS, IRS/CIRS, and T cell growth profiles between patients with aMCI and controls. All differences remained significant after FDR p correction (at p=0.0468). The estimated marginal mean values of the biomarkers acquired from the GLM analysis, as demonstrated in Table 2, are presented in ESF, Table 3. Healthy controls had higher levels of M1, Th1, Th2, IRS, CIRS, IRS/CIRS, Chemokine, T cell growth, and PC_Immune scores compared to subjects with aMCI.

Since the primary analysis on the immune profiles yielded significant results, we have examined the differences in the separate cytokines, chemokines, and growth factors between the study groups. The aMCI group had lower levels of IL1β, IL6, IL7, IL12p70, IL13, GM-CSF, and MCP-1 in comparison to controls (see Table 2 and ESF, Table 3). As reviewed in the Discussion, these 7 cytokines/chemokines show, at physiologic levels, neuroprotective effects. Consequently, we have computed a z unit-based composite as IL1β (z IL1β) + z IL6 + z IL7 + z IL12p70 + z IL13 + z GM-CSF + z MCP1, labelled neuroprotective index or 7NP.

Consequently, we have also computed the differences between the mCoDy group and controls as shown in ESF, Tables 4 and 5. The healthy control group exhibited higher mean values for the Th1, IRS, T cell growth, and PC_Immune profiles, in comparison to the mCoDy group. The series mean values of various pro-inflammatory cytokines, including IL1β, IL6, IL7, IL12p70, GM-CSF, and MCP-1, were found to be greater in the healthy control group compared to the mCoDy group.

### Differences in neurotoxicity and neuroprotection scores between the diagnostic groups

Our results of multivariable regression analyses showed that 3 chemokines frequently appeared as explanatory variables after considering the effects of the 7NP index, namely CXCL8, CCL5, and CCL11. Consequently, we have computed an index of neurotoxic chemokines as z CXCL8 + z CCL11 + z CCL5, denoted as neurotoxic (3NT) index (the Discussion reviews their NT potential). Consequently, we computed the z 3NT – z 7NP composite (labeled 3NT/7NP). **Table 3** shows the mean values of 3NT, 7NP, and 3NT/7NP in aMCI and control subjects. There was no statistically significant difference in 3NT between aMCI and controls, whereas 7NP was significantly lowered in aMCI as compared with controls.

### Prediction of aMCI using immune markers

**Table 4** presents the results of a binary logistic regression analysis conducted to investigate whether immune markers predict aMCI and mCoDy versus controls. The dependent variable in this analysis was the presence of aMCI or mCoDy with controls as the reference group. The results of Model 1 demonstrated that education years, 7NP (both inversely), and 3NT (positively) had a substantial impact on the probability of developing aMCI (effect size of 0.381). Model 2 used the separate cytokines/chemokines in the same framework. The effect size of this model was shown to be higher (Nagelkerke increased to 0.551). The best predictors of aMCI were IL7, IL1β, MCP-1, CCL5, and education. Removing CCL5 showed that CXCL8 became a significant predictor (p=0.005). In Model 3, the restricted mCoDy group was utilized as dependent variable (and controls as reference group). The model showed a better fit comparable to Model 1 in terms of its effect size (Nagelkerke pseudo-R^2^ = 0.447), while using the same predictors.

### Correlation matrix between cognitive test results and immune profiles

As presented in **Table 5**, the qMCI score showed significant and inverse associations with 7NP, IL1β, IL6, IL7, IL12p70, IL13, GM-CSF, and MCP-1, and a positive correlation with the 3NT/7NP ratio. The same variables were also associated with the MoCa score (but all in the opposite direction as compared with the qMCI score). The MMSE score was significantly associated with the 3NT/7NA ratio (inversely), IL6, IL13, and MCP-1 (all positively). The study findings indicate that there is no significant correlation between DSOA scores and any of the cognitive tests and immune markers.

### Prediction of the qMCI and DSOA scores using immune markers

**Table 6** shows the outcome of regression analyses with the qMCI and DSOA scores as dependent variables and either the immune profiles or the separate cytokine/chemokines as explanatory variables. Model 1 shows that education, 3NT, PC_Immune, and age largely predict the qMCI score (32.8% of the variance). Thus, lowered immune functions, higher neurotoxicity and age, and lower education were associated with higher qMCI scores. Model 2 shows that CCL5 (positively) and education, MCP-1, IL13, IL7, and IL-1β (all inversely) were significantly associated with qMCI and explained 38.2% of its variance. Model 3 shows that 3NT and 7NP did not predict the DSOA score. In addition, after using all immune profiles or cytokines/chemokines/growth factors as explanatory variables, it was found that none of the immune variables or any of their combinations could explain the DSOA score.

## Discussion

### Immune-inflammatory biomarkers in aMCI

The first major finding of our study is the notable assocation between the diagnosis of aMCI and immune-inflammatory profiles. Notably, there were significant reductions in Th1, IRS, IRS/CIRS, T cell growth, and PC_immune profiles in individuals with aMCI compared to healthy controls. Our findings indicate that aMCI is distinguished by a suppression of specific immune functions and a general immunosuppression, rather than immune activation or inflammation. Furthermore, significant reductions were observed in IL1β, IL6, IL7, IL12p70, IL13, GM-CSF, and MCP-1 levels in individuals with aMCI compared to the control group.

Therefore, these results contradict previous findings in MCI. As previously discussed in the Introduction, several studies have reported elevated levels of inflammatory or immune markers in individuals with MCI (Contreras et al., 2022; Schmidt-Morgenroth et al., 2023; Shen et al., 2018; Zheng et al., 2019). In the current study, it was found that two out of the three cytokines/chemokines analyzed by Shen et al. (2018) showed a decrease rather than an increase. Specifically, IL6 and MCP-1 exhibited a decrease in their levels in aMCI. As such, our results agree with those of a previous meta-analysis which found no significant variations in immune markers between individuals with MCI and control subjects (Saleem et al., 2015).

One of the main reasons for these variations is that our study was conducted on a highly selected and well-phenotyped group of individuals with aMCI or mCoDy, who were meticulously chosen to exclude those with MDD and DSOA symptoms. Including MDD patients in the aMCI study group can introduce bias due to the characteristic increase in M1, Th1, IRS, T cell growth, and chemokine profiles observed in MDD (Maes & Carvalho, 2018). In addition, the current study took into account BMI, age, and sex when analyzing the data. These factors can potentially introduce bias in immune variables.

### Lowered levels of neuroprotective cytokines/chemokines in aMCI

It is worth mentioning that the cytokines/chemokines that are downregulated in aMCI have pro-inflammatory effects when increased, as observed in MDD (Maes and Carvalho, 2018). However, it should be stressed that the 7 cytokines/chemokines that exhibit a decrease in aMCI have been found to have neuroprotective effects at normal levels. These include IL1β, IL6, IL7, IL12p70, IL13, GM-CSF, and MCP-1. As an illustration, there is empirical evidence suggesting that IL1β, at normal physiological levels, may play a role in maintaining the health and functioning of neurons (Alsbrook et al., 2023; Poh et al., 2022). In a recent study conducted by Alsbrook et al. (2023), it was found that IL1β plays a crucial role in the intricate processes of tissue healing and neuroplasticity. Excessive levels of IL6 are usually linked to negative effects that can lead to cognitive impairment (Maes et al., 2014). However, maintaining normal levels of IL6 seems to be crucial for preserving different aspects of neuronal health (Chae et al., 2016; Maes et al., 2014). Research has shown that IL7 plays a crucial role in maintaining glucose homeostasis and has the potential to aid in neurorecovery following brain damage (Wofford et al., 2008). IL12p70 plays a crucial role in preserving neuronal health by regulating the inflammatory conditions in the central nervous system (Verma et al., 2014). In a recent study by Huan et al. (2022), it was found that IL12p70 plays a crucial role in promoting neurogenesis, synaptic plasticity, regulating neurotrophic factors, and ensuring the survival of neurons. In a recent study conducted by Yang et al. (2022), it was discovered that individuals with higher levels of IL12p70 experience a decelerated cognitive decline. Additionally, these individuals also exhibited reduced tau levels and a decrease in neurodegeneration, particularly among those with elevated amyloid beta. IL13 has been found to play a role in reducing neuroinflammation by promoting the M2 microglia phenotype and contributing to the death of the microglia M1 phenotype, as demonstrated in studies by Mori et al. (2016) and Miao et al. (2020). In addition, IL13 has been shown to enhance functional recovery following traumatic brain injury through its anti-inflammatory and neuroprotective properties (Miao et al., 2020). GM-CSF has the ability to regulate the activities of microglia, which can potentially improve the survival of neurons (Dikmen et al., 2020). This suggests that GM-CSF plays a role in maintaining the health of neurons when present at normal concentrations (Dikmen et al., 2020). In addition, this factor activates mesenchymal stem cells (MSCs) found in the bone marrow. MSCs have been studied for their positive impact on cognition and regeneration, particularly in relation to Alzheimer’s disease (Hernández & García, 2021; Manczak et al., 2009) and impaired neurocognitive functions (Nakazaki et al., 2019). The potential role of MCP-1 in mediating the neuroprotective effects of noradrenaline and its ability to prevent ATP loss has been suggested by Madrigal et al. (2009).

The reduced levels of these cytokines and the general suppression of the immune system suggest that aMCI and mCoDy are not caused by heightened immune activation or inflammation. Our results suggest that the protective functions of these cytokines/chemokines, which are typically achieved at normal levels, may be weakened in subjects with aMCI, potentially leading to reduced neuroprotection.

### Neurotoxic cytokines/chemokines in aMCI and mCoDy

It is worth noting that our multivariate regression analyses revealed an intriguing finding. Upon incorporating the aforementioned neuroprotective immune compounds, three neurotoxic chemokines, specifically CCL11, CCL5, and CXCL8, emerged as significant predictors in the final regression model. By analyzing a combination of these three chemokines, we were able to create a composite score. This score allowed us to determine that a decrease in the neuroprotective index, along with an increase in this neurotoxicity index, strongly predicted aMCI or mCoDy as compared to controls. CCL5 (or RANTES) has been associated with a wide variety of neurodegenerative diseases (Tripathy et al., 2010). CCL11 plays a significant role in elucidating the cognitive impairments associated with depression and major psychoses. It disrupts neurogenesis and the functions of the BBB, as reviewed by Ivanovska et al. (2020). CXCL8 plays a crucial role in the intricate web of inflammation and may be linked to cognitive impairment and various mental health conditions (Tsai, 2021).

On the other hand, other neurotoxic cytokines such as IL15 and IL17 were not even measurable in the current study group. According to a study conducted by Di Castro et al. (2023), IL15, a proinflammatory cytokine, has been found to have a negative impact on episodic memory. IL17 plays a significant role in promoting inflammation, disrupting the blood-brain barrier (BBB), and causing tissue damage in conditions such as Alzheimer’s disease (Milovanovic et al., 2020). Research has shown that IL17 has been linked to negative effects on neuronal functions and overall brain health (Lu et al., 2023). Nevertheless, these cytokines, which play a role in MDD (Maes and Carvalho, 2018), are not involved in the pathophysiology of aMCI.

### Prediction of the quantitative qMCI and DSOA scores

One notable discovery from this study is the significant correlation between the qMCI score and indicators of immunosuppression and increased neurotoxicity (after considering the effects of confounders such as age and education). These factors, when considered together, accounted for 32.8% of the variance in qMCI. This suggests that aMCI may be influenced by reduced neuroprotection and weakened immune defenses, as indicated by lower levels of certain cytokines and chemokines. The decreased levels of these compounds in aMCI may result in reduced neuroprotection, potentially rendering these individuals more vulnerable to the effects of neurotoxic chemokines. This combination could ultimately lead to neuronal damage and cognitive decline. Therefore, it appears that the delicate interplay between neurotoxic and neuroprotective factors may play a crucial role in the advancement of cognitive decline.

This condition exhibits similarities to recent discoveries in schizophrenia, where a decline in overall cognitive abilities, including executive functions, attention, semantic and episodic memory, is linked to an elevated ratio of immune-linked neurotoxicity to neuroprotection (Maes, 2023; Maes et al., 2023). However, it is worth noting that the neurotoxicity index shows a significant increase in schizophrenia, which helps to explain why the neurocognitive disorders in deficit schizophrenia are more pronounced compared to aMCI (Kanchanatawan et al., 2018).

Nevertheless, the study findings suggest that there are no significant associations between the DSOA symptoms and the immune markers that were examined. In a previous study, it was discovered that a significant portion of the variation in the DSOA score could be attributed to psychosocial stressors, specifically negative life events and adverse childhood experiences (Tran-Chi et al., 2024). It appears that DSOA, as opposed to the more severe depressive symptoms of MDD, is characterized by a psychological state in which negative cognitions dominate. On the other hand, in MDD, the heightened neurotoxicity associated with the immune system is influenced, to some extent, by negative experiences during childhood and adverse events in life (Almulla et al., 2024).

## Conclusions

Our thorough investigation provides insight into the various dynamics of aMCI, highlighting the significant impact of immunosuppression and reduced neuroprotection, as well as the elevated neurotoxicity of CCL11, CCL5, and CXCL8. The findings emphasize the importance of maintaining a delicate balance between neurotoxicity and neuroprotection during the progression of aMCI. Immunosuppression seems to be the main factor contributing to aMCI, rather than immune activation. It is possible to consider that the decline in immune function and reduced neuroprotection could be a contributing factor in the progression from mild cognitive impairment to Alzheimer’s disease. Our findings deserve replication in other countries and ethnicities. If replicated, the immunosuppression of neuroprotective cytokines/chemokines may be a new drug target to treat aMCI. Future research on aMCI should always consider MDD and the possible intervening effects of DSOA symptoms.

## Availability of data and materials

The dataset generated during and/or analysed during the current study will be available from MM upon reasonable request and once the authors have fully exploited the dataset.

## Author’s contributions

All authors contributed to the paper. MM, CT, and V-LT-C: conceptualization and study design. V-LT-C and MM: first draft writing. V-LT-C, MM, CT, SH, GN, and MS: editing. GN, CT and V-LT-C: recruitment of patients. MM and V-LT-C: Statistical analyses. All authors revised and approved the final draft.

## Institutional Review Board Statement

This study was approved by the Institutional Review Board (IRB) of the Faculty of Medicine, Chulalongkorn University, Bangkok, Thailand (IRB no. 0372/65), which complies with the International Guideline for Human Research Protection as required by the Declaration of Helsinki.

## Informed consent

Before taking part in the study, all participants and/or their caregivers provided written informed consent.

## Supporting information

ESF

