## Supplementary material for "Is amnestic mild cognitive impairment a neuro-immune condition?": Tables

**Table 1.** Socio-demographic data of healthy controls (HC), and subjects with amnestic mild cognitive impairment (aMCI)

| **Clinical Variables** | **HC (n=60)** | **aMCI (n=61)** | ***F/ χ^2^*** | ***df*** | ***p*** |
| --- | --- | --- | --- | --- | --- |
| Age – years (±SD) | 66.80 (±4.07) | 68.72 (±3.83) | 7.134 | 1/119 | 0.009 ^a^ |
| Sex (F/M) | 47/13 | 44/17 | 0.624 | 1 | 0.430 ^b^ |
| Education – years (±SD) | 15.88 (±2.94) | 13.70 (±4.46) | 10.019 | 1/119 | 0.002 ^a^ |
| Body mass index (±SD) (kg/m^2^) | 22.66 (±2.92) | 23.26 (±3.50) | 1.072 | 1/119 | 0.303 ^a^ |
| Single/married/divorced, separated or widowed | 15/36/9 | 14/40/7 | 0.487 | 2 | 0.784 ^b^ |
| MMSE (±SD) score | 28.70 (±1.23) | 26.39 (±2.27) | 47.737 | 1/119 | <0.001 ^a^ |
| MoCA (±SD) score | 27.43 (±1.51) | 22.33 (±1.65) | 314.65 | 1/119 | <0.001 ^a^ |
| qMCI (±SD) score | -0.83 (±0.44) | 0.82 (±0.65) | 266.53 | 1/119 | <0.001 ^a^ |
| DSOA (±SD) score | -0.18 (±0.98) | 0.18 (±0.99) | 4.025 | 1/119 | 0.047 ^a^ |

*Note:* SD, Standard Deviation; ^a^ One-way ANOVA; ^b^ χ^2^ – test; F/ χ^2^, Results of analyses of variance (F); ^b^ analyses of contingency analyses (χ^2^); df, degree of freedom; p, p value.

*Abbreviations*: MMSE, Mini-Mental State Examination; MoCA, Montreal Cognitive Assessment; qMCI, quantitative Mild Cognitive Impairment score; DSOA, Distress Symptoms of Old Age score.

**Table 2.** Results of General Linear Model analyses that examine the association between immune-inflammatory biomarkers and the diagnosis of amnestic mild cognitive impairment (aMCI) versus healthy controls (HC), while adjusting for age, sex, and body mass index.

| **Tests** | **Dependent Variables** | **Explanatory variables** | ***F*** | ***df*** | ***p*** | **Partial Eta Squared** |
| --- | --- | --- | --- | --- | --- | --- |
| **Univariate GLM** | M1  Th1  Th2  IRS  CIRS  IRS/CIRS  Chemokine  T cell growth  PC_Immune | aMCI/HC  aMCI/HC  aMCI/HC  aMCI/HC  aMCI/HC  aMCI/HC  aMCI/HC  aMCI/HC  aMCI/HC | 3.246  9.818  2.144  9.815  2.372  5.079  1.455  10.493  8.236 | 1/119  1/119  1/119  1/119  1/119  1/119  1/119  1/119  1/119 | 0.074  0.002  0.146  0.002  0.126  0.026  0.230  0.002  0.005 | 0.027  0.076  0.018  0.076  0.020  0.041  0.012  0.081  0.065 |
| **Significant Univariate**  **GLM** | IL1β  IL6  IL7  IL12p70  IL13  GM-CSF  MCP-1 | aMCI/HC  aMCI/HC  aMCI/HC  aMCI/HC  aMCI/HC  aMCI/HC  aMCI/HC | 13.902  4.371  7.941  10.685  4.534  7.830  6.626 | 1/119  1/119  1/119  1/119  1/119  1/119  1/119 | <0.001  0.039  0.006  0.001  0.035  0.006  0.011 | 0.105  0.035  0.063  0.082  0.037  0.062  0.053 |

*Note:* F, Results of GLM analysis; df, degree of freedom; p, p value.

*Abbreviations*: IL, Interleukin; IRS/CIRS, Immune-Inflammatory Response System/Compensatory Immunoregulatory System; Th, T helper; GM-CSF, Granulocyte-Macrophage Colony Stimulating Factor; MCP, Monocyte Chemoattractant Protein; GF, Growth Factor; PC_Immune, a principal component extracted from the macrophage M1; Th1 (T helper), Th2, immune-inflammatory responses system (IRS) and the compensatory immunoregulatory system (CIRS). See ESF, Table 1 and 2 for explanation and computation.

**Table 3.** Results of multivariate General Linear Model analyses which examine the associations between the Neurotoxic and Neuroprotective immune indices and their ratio and the diagnosis of amnestic mild cognitive impairment (aMCI) versus healthy controls (HC).

| **Variables** | **HC (n=60)** | **aMCI (n=61)** | ***F*** | ***df*** | ***p*** |
| --- | --- | --- | --- | --- | --- |
| Neurotoxic (3NT) index | -0.030 (0.130) | 0.030 (0.129) | 0.107 | 1/119 | 0.744 |
| Neuroprotective (7NP) index | 0.342 (0.122) | -0.337 (0.121) | 15.636 | 1/119 | <0.001 |
| 3NT/7NP | -0.364 (0.121) | 0.358 (0.120) | 18.005 | 1/119 | <0.001 |

*Note:* F, Results of analyses of variance; df, degree of freedom; p, p value.

**Table 4.** Results of binary logistic regression with diagnosis as dependent variable and the immune biomarkers and solitary cytokines and chemokines as explanatory variables

| **Dependent**  **variables** | **Explanatory variable** | **B** | **S.E.** | **W** | **P** | **OR** | **95% CI** | **Omnibus model X²** | **df** | **p** | **Nagelkerke pseudo-R2** |
| --- | --- | --- | --- | --- | --- | --- | --- | --- | --- | --- | --- |
| Model 1 | | | | | | | | 40.745 | 3 | <0.001 | 0.381 |
|  | Education | -0.245 | 0.07 | 12.21 | <0.001 | 0.783 | 0.683-0.898 |  | | | |
|  | 7NP | -1.682 | 0.398 | 17.845 | <0.001 | 0.186 | 0.085-0.406 |  |  |  |  |
|  | 3NT | 0.386 | 0.12 | 10.43 | 0.001 | 1.472 | 1.164-1.861 |  |  |  |  |
| Model 2 | | | | | | | | 64.507 | 5 | <0.001 | 0.551 |
|  | Education | -0.237 | 0.074 | 10.356 | 0.001 | 0.789 | 0.683-0.911 |  | | | |
|  | IL1β | -1.065 | 0.271 | 15.393 | <0.001 | 0.345 | 0.203-0.587 |  |  |  |  |
|  | IL7 | -1.257 | 0.315 | 15.913 | <0.001 | 0.284 | 0.153-0.528 |  |  |  |  |
|  | MCP-1 | -1.067 | 0.302 | 12.518 | <0.001 | 0.344 | 0.190-0.621 |  |  |  |  |
|  | CCL5 | 1.084 | 0.335 | 10.448 | 0.001 | 2.956 | 1.532-5.702 |  |  |  |  |
| Model 3 | | | | | | | | 44.500 | 3 | <0.001 | 0.447 |
|  | Education | -0.35 | 0.086 | 16.492 | <0.001 | 0.705 | 0.595-0.834 |  | | | |
|  | 7NP | 1.045 | 0.304 | 11.795 | 0.001 | 2.843 | 1.566-5.161 |  |  |  |  |
|  | 3NT | -1.562 | 0.407 | 14.7 | <0.001 | 0.21 | 0.094-0.466 |  |  |  |  |

*Note:* S.E., Standard Error of the Coefficient; W, Wald; OR, Odds ratio; CI, Confidence Intervals; χ^2^, Chi-square tests; df, degree of freedom; p, p value.

*Abbreviations*: NT, immune-linked neurotoxicity index; NP, attenuated immune-linked neuroprotection index; IL, interleukin; MCP, Monocyte Chemoattractant Protein; CCL, C-C motif chemokine ligand.

**Table 5**. Correlation matrix between clinical and neuropsychiatric scores with the measured immune markers

|  | **qMCI** | **DSOA** | **MoCA** | **MMSE** |
| --- | --- | --- | --- | --- |
| qMCI | 1 | 0.138 | -0.901^**^ | -0.860^**^ |
| DSOA | 0.138 | 1 | -0.165 | -0.060 |
| MoCA | -0.901^**^ | -0.165 | 1 | 0.561^**^ |
| MMSE | -0.860^**^ | -0.060 | 0.561^**^ | 1 |
| 3NT | 0.062 | 0.121 | -0.051 | -0.063 |
| 7NP | -0.232^*^ | -0.056 | 0.216^*^ | 0.156 |
| 3NT/7NP | 0.287^**^ | 0.172 | -0.261^**^ | -0.214^*^ |
| IL1β | -0.224^*^ | -0.004 | 0.250^**^ | 0.093 |
| IL6 | -0.327^**^ | 0.060 | 0.257^**^ | 0.299^**^ |
| IL7 | -0.249^**^ | -0.098 | 0.249^**^ | 0.154 |
| IL12p70 | -0.242^**^ | -0.146 | 0.244^**^ | 0.147 |
| IL13 | -0.253^**^ | -0.052 | 0.198^*^ | 0.246^**^ |
| GM-CSF | -0.250^**^ | -0.042 | 0.273^**^ | 0.137 |
| MCP-1 | -0.289^**^ | -0.027 | 0.239^**^ | 0.252^**^ |

*Note:* *, p value < 0.05; **, p value < 0.01.

*Abbreviations*: qMCI, quantitative Mild Cognitive Impairment score; DSOA, Distress Symptoms of Old Age score; MoCA, Montreal Cognitive Assessment; MMSE, Mini-Mental State Examination; NT, immune-linked neurotoxicity index; NP, attenuated immune-linked neuroprotection index; IL, Interleukin; GM-CSF, Granulocyte-Macrophage Colony Stimulating Factor; MCP, Monocyte Chemoattractant Protein.

**Table 6**. Results of multiple regression analyses with quantitative Mild Cognitive Impairment (qMCI) scores as dependent variables

| **Dependent variables** | **Explanatory variable** | **Coefficients of input variables** | | | **Model statistics** | | | |
| --- | --- | --- | --- | --- | --- | --- | --- | --- |
|  |  | **β** | **t** | **p** | **F** | **df** | **p** | **R^2^** |
| **#1. qMCI** | **Model** | | | | 14.163 | 4/116 | <0.001 | 0.328 |
|  | Education | -0.366 | -4.571 | <0.001 |  |  |  |  |
|  | 3NT | 0.585 | 4.820 | <0.001 |  |  |  |  |
|  | PC_Immune | -0.582 | -4.809 | <0.001 |  |  |  |  |
|  | Age | 0.175 | 2.187 | 0.031 |  |  |  |  |
| **#2. qMCI** | **Model** | | | | 11.753 | 6/114 | <0.001 | 0.382 |
|  | Education | -0.392 | -5.260 | <0.001 |  |  |  |  |
|  | MCP-1 | -0.309 | -3.719 | <0.001 |  |  |  |  |
|  | IL13 | -0.201 | -2.553 | 0.012 |  |  |  |  |
|  | CCL5 | 0.338 | 3.913 | <0.001 |  |  |  |  |
|  | IL7 | -0.226 | -2.759 | 0.007 |  |  |  |  |
|  | IL1β | -0.149 | -1.995 | 0.048 |  |  |  |  |
| **#3. DSOA** | **Model** | | | | 1.622 | 1.816 | 0.148 | 0.045 |
|  | Education | 0.120 | 1.302 | 0.196 |  |  |  |  |
|  | 3NT | 0.169 | 1.611 | 0.110 |  |  |  |  |
|  | NP | -0.134 | -1.286 | 0.201 |  |  |  |  |

*Note:* #, number of regression analysis; β, Standardized regression coefficient; t, t-statistic value; R^2^, Total variance explained; F, Results of analyses of variance; df, degree of freedom; p, p-value.

*Abbreviations*: NT, immune-linked neurotoxicity index; PC_Immune, a principal component extracted from immune variables - first principal component extracted from macrophage M1, Th-1 (T-helper), Th-2, and the immune-inflammatory responses system. Fr cytokines: see ESF, Table 1.
