## Supplementary material for "Is amnestic mild cognitive impairment a neuro-immune condition?": ESF

**ELECTRONIC SUPPLEMENTARY FILE (ESF)**

**ESF, Table 1**. Overview of the cytokines, chemokines, and growth factors measured in the current study.

| **Protein abbreviations** | **Gene Symbol** | **> OOR (%)** | **Protein name / alias** |
| --- | --- | --- | --- |
| **IFN-γ** | IFNG | 100 | Interferon-γ |
| **IL1β** | IL1B | 85 | Interleukin-1β |
| **sIL1RA** | IL1RN | 68.3 | Soluble interleukin-1 receptor antagonist |
| **IL2** | IL2 | 0 | Interleukin-2 |
| **IL4** | IL4 | 100 | Interleukin-4 |
| **IL5** | IL5 | 8.3 | Interleukin-5 |
| **IL6** | IL6 | 68.3 | Interleukin-6 |
| **IL7** | IL7 | 58.3 | Interleukin-7 |
| **IL9** | IL9 | 100 | Interleukin-9 |
| **IL10** | IL10 | 51.3 | Interleukin-10 |
| **IL12p70** | IL12RB1 | 72.5 | Interleukin-12 p70 |
| **IL13** | IL13 | 51.7 | Interleukin-13 |
| **IL15** | IL15 | 0 | Interleukin-15 |
| **IL17** | IL17A | 0 | Interleukin-17 |
| **TNF-α** | TNF | 100 | Tumor necrosis factor-α |
| **G-CSF** | CSF3 | 100 | Granulocyte colony stimulating factor (G-CSF) or colony stimulating factor 3 (CSF3) |
| **GM-CSF** | CSF2 | 57.5 | Granulocyte-macrophage colony-stimulating factor (GM-CSF) or colony-stimulating factor 2 (CSF2) |
| **CCL2 or MCP1** | CCL2 | 100 | C-C motif chemokine ligand 2 (CCL2) or monocyte chemoattractant protein 1 (MCP1) |
| **CCL3 or MIP-1α** | CCL3 | 100 | C-C motif Chemokine ligand 3 (CCL3) or macrophage inflammatory protein 1-alpha (MIP-1α) |
| **CCL4 or MIP-1β** | CCL4 | 100 | C-C motif chemokine ligand 4 (CCL4) or macrophage inflammatory protein 1β (MIP-1β) or lymphocyte activation gene 1 protein |
| **CCL5 or RANTES** | CCL5 | 100 | C-C motif chemokine ligand 5 (CCL5) or regulated upon activation, normally T-expressed, and presumably Secreted (RANTES) |
| **CCL11 or Eotaxin** | CCL11 | 100 | C-C motif chemokine ligand 11 (CCL11) or eosinophil chemotactic protein |
| **CXCL8 or IL8** | CXCL8 | 100 | C-X-C motif chemokine ligand 8 (CXCL8) or interleukin-8 (IL-8) |
| **CXCL10 or IP10** | CXCL10 | 100 | C-X-C motif chemokine ligand 10 (CXCL10) or Interferon gamma-induced protein 10 (IP10) |
| **FGF** | FGF2 | 100 | Fibroblast growth factor 2 (FGF) or basic fibroblast growth factor |
| **PDGF** | PDGFA | 100 | Platelet derived growth factor (PDGF) |
| **VEGF** | VEGFA | 7.5 | Vascular endothelial growth factor (VEGF) |

Adapted from: *Maes M, Rachayon M, Jirakran K, Sodsai P, Klinchanhom S, Gałecki P, Sughondhabirom A, Basta-Kaim A. The Immune Profile of Major Dysmood Disorder: Proof of Concept and Mechanism Using the Precision Nomothetic Psychiatry Approach. Cells. 2022 Mar 31;11(7):1183. doi: 10.3390/cells11071183. PMID: 35406747; PMCID: PMC8997660.*

*Kalayasiri R, Dadwat K, Supaksorn T, Sirivichayakul S, Maes M. Methamphetamine (MA) use, MA dependence, and MA-induced psychosis are associated with increasing aberrations in the compensatory immunoregulatory system and interleukin-1α and CCL5 levels. medRxiv 2023.03.26.23287766; doi: https://doi.org/10.1101/2023.03.26.23287766*

**ESF, Table 2**. Construction of the various immune profiles in this study

| **Immune Profile** | **Members** |
| --- | --- |
| **M1 macrophage** | IL1β, IL6, TNF-α, CXCL8, CCL3 |
| **T helper-1** | IFN-γ, IL12p70 |
| **T helper-2** | IL4, IL9, IL13 |
| **IRS** | IL1β, IL6, TNF-α, CXCL8, CCL3, IFN-γ, IL12p70, G-CSF, GM-CSF, CXCL10, CCL5, CCL2 |
| **CIRS** | IL4, IL10, sIL1RA |
| **IRS/CIRS** | z IRS – z CIRS |
| **Chemokines** | CCL2, CCL3, CCL4, CCL5, CCL11, CXCL8, CXCL10 |
| **T cell growth** | IL4, IL9, IL12p70, GM-CSF |
| **PC_Immune** | The first PC extracted from M1. Th1, Th2, IRS, and CIRS. |

IRS: immune-inflammatory response system; CIRS: compensatory immunoregulatory system

*Adapted from: Rachayon, M.; Jirakran, K; Sodsai, P; Klinchanhom, S.; Sughondhabirom, A.; Plaimas, K.; Suratanee, A.; Maes, M. Effects of cannabidiol on activated immune-inflammatory pathways in major depressive patients and healthy controls. MedRxiv 2022.02.04.22270489; doi:https://doi. org/10.1101/2022.02.04.22270489*

**ESF, Table 3**. Model-generated estimated marginal mean (SE) values (expressed as z-scores and series mean) obtained by multivariate GLM analysis with HC and aMCI as groups.

| **Immune profiles** | **HC**  ***n = 60*** | **aMCI**  ***n = 61*** |
| --- | --- | --- |
| M1  Th1  Th2  IRS  CIRS  IRS/CIRS  Chemokine  T cell growth  PC_Immune  IL1β  IL6  IL7  IL12p70  IL13  GM-CSF  MCP-1 | 0.164 (0.128)  0.278 (0.125)  0.134 (0.129)  0.278 (0.125)  0.141 (0.129)  0.204 (0.127)  0.111 (0.129)  0.287 (0.125)  0.257 (0.126)  0.885 (0.009)  0.456 (0.038)  2.383 (0.118)  0.965 (0.127)  1.352 (0.162)  2.668 (0.237)  1.642 (0.04) | -0.162 (0.127)  -0.274 (0.124)  -0.132 (0.128)  -0.274 (0.124)  -0.139 (0.128)  -0.201 (0.126)  -0.109 (0.128)  -0.282 (0.124)  -0.252 (0.125)  0.836 (0.009)  0.345 (0.037)  1.914 (0.117)  0.439 (0.126)  0.867 (0.16)  1.733 (0.235)  1.496 (0.04) |

*Abbreviations*: aMCI, subjects with amnestic Mild Cognitive Impairment; HC, Healthy Controls; IL, Interleukin; IRS/CIRS, Immune-Inflammatory Response System/Compensatory Immunoregulatory System; Th, T helper; GM-CSF, Granulocyte-Macrophage Colony Stimulating Factor; MCP, Monocyte Chemoattractant Protein; PC_Immune, a principal component extracted from immune variables - first principal component extracted from macrophage M1, Th-1 (T-helper), Th-2, immune-inflammatory responses system.

**ESF, Tabe 4.** Multivariate General Linear Model analyses that examine the association between immune-inflammatory biomarkers and the diagnosis of amnestic mild cognitive dysfunction (mCoDy) versus healthy controls (HC)

| **Tests** | **Immune Variables** | **Diagnosis** | ***F*** | ***df*** | ***p*** | **Partial Eta Squared** |
| --- | --- | --- | --- | --- | --- | --- |
| Significant Univariate | Th1  IRS  T cell growth  PC_Immune  IL1β  IL6  IL7  IL12p70  GM-CSF  MCP-1 | mCoDy/HC  mCoDy/HC  mCoDy/HC  mCoDy/HC  mCoDy/HC  mCoDy/HC  mCoDy/HC  mCoDy/HC  mCoDy/HC  mCoDy/HC | 4.996  5.019  5.226  4.193  8.143  5.329  6.663  5.710  4.721  4.448 | 1/107  1/107  1/107  1/107  1/107  1/107  1/107  1/107  1/107  1/107 | 0.027  0.027  0.024  0.043  0.005  0.023  0.011  0.019  0.032  0.037 | 0.045  0.045  0.047  0.038  0.071  0.047  0.059  0.051  0.042  0.040 |

*Note:* F, Results of analyses of variance (F); df, degree of freedom; p, p value.

*Abbreviations*: mCoDy, subjects with Mild Cognitive Dysfunction; HC, Healthy Controls; IL, Interleukin; IRS/CIRS, Immune-Inflammatory Response System/Compensatory Immunoregulatory System; Th, T helper; GM-CSF, Granulocyte-Macrophage Colony Stimulating Factor; MCP, Monocyte Chemoattractant Protein.

**ESF, Table 5**. Model-generated estimated marginal mean (SE) values (expressed as z-scores and series mean) obtained by multivariate GLM analysis with HC and mCoDy as groups.

| **Biomarkers** | **HC**  ***n = 37*** | **mCoDy**  ***n = 52*** |
| --- | --- | --- |
| Th1  IRS  T cell growth  PC_Immune  IL1β  IL6  IL7  IL12p70  GM-CSF  MCP-1 | 0.264(0.133)  0.269(0.133)  0.277(0.132)  0.249(0.133)  0.886(0.009)  0.447(0.037)  2.375(0.125)  0.94(0.137)  2.64(0.253)  1.646(0.043) | -0.167(0.139)  -0.164(0.14)  -0.161(0.139)  -0.146(0.14)  0.847(0.01)  0.322(0.039)  1.907(0.131)  0.512(0.143)  1.844(0.265)  1.515(0.045) |

*Abbreviations*: mCoDy, subjects with Mild Cognitive Dysfunction; HC, Healthy Controls; IL, Interleukin; IRS/CIRS, Immune-Inflammatory Response System/Compensatory Immunoregulatory System; Th, T helper; GM-CSF, Granulocyte-Macrophage Colony Stimulating Factor; MCP, Monocyte Chemoattractant Protein; PC_Immune, a principal component extracted from immune variables - first principal component extracted from macrophage M1, Th1 (T helper), Th2 and IRS.
